# Stepwise explorative model to determine pathogenicity of cultured blood isolates

**DOI:** 10.1101/2025.10.24.25338513

**Authors:** Shiv Narayan Sahu, Balram Ji Omar, Mukesh Bairwa, Prakhar Sharma, Partha Sahu, Monika Saran, Prasan Kumar Panda

## Abstract

Blood culture remains the gold standard for diagnosing bloodstream infections (BSIs); however, distinguishing true pathogens from contaminants remains a critical challenge. Misclassification can lead to inappropriate antimicrobial use, prolonged hospitalization, and increased healthcare burden. This study proposes a structured, stepwise clinical-pathological model to differentiate pathogenic from non-pathogenic organisms in blood cultures.

In this prospective study at a tertiary hospital in northern India, 205 blood culture-positive adults were evaluated between August and October 2024. Organisms were classified as pathogenic or non-pathogenic using microbiological data and clinical indicators, including SOFA score, time to positivity, and site concordance, through a seven-step algorithm with 28-day outcome follow-up.

Among 1600 blood culture samples received, 205 isolates were positive for an organism, 160 (78.0%) were identified as pathogenic and 45 (22.0%) as non-pathogenic. The most common pathogens were *Klebsiella pneumoniae* (20.0%), *Acinetobacter baumannii* (9.3%), and *Pseudomonas aeruginosa* (6.3%), while non-pathogens were mainly coagulase-negative staphylococci (CONS, 18.5%) and *Stenotrophomonas maltophilia* (8.3%). Mean Time to Positivity (TTP) was significantly shorter in pathogens (16.3 ± 8.0 hours) compared to non-pathogens (21.5 ± 10.1 hours; p < 0.001). Discordance was observed in 7 cases (3.4%) where clinicians labelled isolates as non-pathogens but microbiologists disagreed, and in 26 cases (12.7%) with the opposite interpretation. Overall agreement was 65.4%, with a Cohen’s kappa of 0.25, indicating fair inter-rater reliability. This stepwise clinical-microbiological model offers an effective framework for distinguishing pathogens from non-pathogens in BSIs. Incorporating SOFA score, TTP, and culture concordance enhances diagnostic stewardship, informs antimicrobial decisions, and supports prognostication, especially in resource-limited and high-burden healthcare settings.

## Introduction

Blood culture remains the cornerstone for diagnosing bloodstream infections (BSIs), enabling identification of causative organisms and guiding targeted antimicrobial therapy. However, contamination rates may range from one-third to one-half, often due to factors such as improper collection, delayed transport, or suboptimal laboratory handling.(1) One of the persistent challenges in clinical microbiology is distinguishing true pathogens from non-pathogenic commensals, colonisers, or contaminants. This differentiation is critical, as over-interpretation may lead to unnecessary investigations, antimicrobial overuse, prolonged hospitalisation, and increased healthcare costs, while under-recognising true pathogen risks may delay or miss treatment.(2)

Despite being a gold standard, blood cultures are sometimes obtained without clear clinical indications, compounding the issue of false positives and non-pathogenic isolates. Laboratory reports rarely delineate the pathogenic significance of an isolate, placing interpretative responsibility variably on the microbiologist or treating clinician, raising the question of whether a structured, collaborative, and algorithmic approach could enhance diagnostic accuracy.(1)

A pathogen is any microbe capable of causing host damage, whether through direct microbial effects or via the host’s immune response. This definition includes both classical pathogens and opportunistic organisms that cause disease under specific conditions.(3) Commensals are organisms that coexist with the host without inducing a pathogenic response. Colonisers persist without causing local damage but may become pathogenic under specific conditions.(4) Contaminants are introduced during sample collection or processing and lack clinical relevance to the patient.(5)

This study proposes a stepwise clinical-pathological model to categorise blood culture isolates as pathogenic or non-pathogenic, integrating microbiological findings with clinical context. The aim is to quantify the proportion of true pathogens and assess the clinical correlates, treatment decisions, and outcomes associated with their classification.

## Methodology

This longitudinal explorative study was conducted to determine the proportion of pathogenic and non-pathogenic organisms in blood culture-positive samples. The study was time bound, took place at a 1000-bedded tertiary care teaching hospital in northern India (AIIMS Rishikesh) and included data collected between August 1, 2024, and October 31, 2024, from the Departments of General Medicine, Nephrology, and Pulmonary Medicine.. Adult patients aged 18 years or older admitted with a positive blood culture report were enrolled in the study. Patients were included if they had a confirmed positive blood culture while admitted to the institution. Exclusion criteria included refusal to participate, inability to follow up, or denial of study inclusion by the treating physician. The study was approved by the Institutional Ethics Committee.

### Data collection

The patients with a positive blood culture were included in the study as per daily visits to the Microbiology department. The reporting microbiologist and treating clinicians were independently asked to classify the identified organism as a pathogen or a non-pathogen, and their responses were recorded. Each isolate was independently assessed by the microbiologist, treating physician, and the investigator for its pathogenic potential, incorporating clinical indicators, SOFA score dynamics, treatment response, and presence of foreign devices. Patients were followed up until discharge or for a maximum of 28 days for the outcome of the patient. The final classification of the organisms was determined by the investigator’s step-wise model prepared on microbiological and clinical evidence, as well as follow-up data.

### Structured stepwise model for organism classification

Whenever a blood culture tested positive and met the inclusion criteria, the organism underwent a seven-step clinical-pathological algorithm to determine its pathogenicity by the investigator team as follows. (supplementary)

- **Step 1:** Evaluate any symptoms and signs of localized infection using Checklist I. → If symptomatic → proceed to Step 2; if asymptomatic □ proceed to Step 3(6)
- **Step 2:** Is the same organism isolated from the local site and blood? If yes, □ classify as Pathogenic, proceed to step 5
- **Step 3:** If a rise in SOFA score ≥ 2 within 48 hours using Checklist II, →proceed to step 4. If no □ proceed to step 7(7)
- **Step 4:** Response to treatment present or any foreign body in the vascular system is the likely source using checklist III □ classify as Pathogenic □proceed to step 5(8)
- **Step 5: C**haracterization of Pathogenic organisms: If any foreign body in the vascular bed is the likely source, □ Pathogenic colonizer
- **Step 6 & 7:** Characterization of Non-Pathogenic organisms
- **Step 6**: Search for other potential sources of sepsis, if no alternative foci □ classify as a non-pathogenic (contaminants)(1)
- **Step 7:** Any foreign body in the vascular system is the likely source □ classify as a non-pathogenic colonizer

The Sequential Organ Failure Assessment (SOFA) score was calculated using standard criteria based on six organ systems: Respiratory, cardiovascular, hepatic, coagulation, renal, and neurological parameters, as per established guidelines.

### Outcome measures

The primary outcome was to determine the proportion of pathogens and non-pathogens in blood cultures using a stepwise model and to identify microbiological and clinical factors associated with pathogenicity.

Once the pathogenicity of the isolated organism was determined, patients were followed prospectively until discharge or up to 28 days—whichever occurred earlier—to evaluate clinical outcomes. Based on the clinical course and treatment decisions, outcomes were classified into four categories: (1) Improved with treatment, applicable to patients with pathogenic organisms who showed clinical recovery following targeted antimicrobial therapy; (2) No response to treatment, representing pathogenic cases where appropriate therapy failed to produce clinical improvement; (3) Not treated, defined for non-pathogenic isolates where no specific antimicrobial therapy was initiated; and (4) Treatment initiated for non-pathogenic organisms. This categorization allowed objective assessment of the appropriateness of pathogen classification and its correlation with patient outcomes.

### Statistical analysis

Data entry was performed using Microsoft Excel, and statistical analysis was conducted using IBM SPSS Statistics for Windows, Version 25.0 (IBM Corp., Armonk, NY, USA). The collected data were analyzed using a combination of descriptive and inferential statistical techniques. Continuous variables were expressed as mean ± standard deviation (SD) or median with interquartile range (IQR), depending on the distribution assessed. The normality of continuous variables was verified using visual and statistical methods, following which either *independent samples t-tests* (for normally distributed variables) or *Wilcoxon-Mann-Whitney U tests* (for non-parametric comparisons) were employed to assess group differences. Categorical variables were summarized as frequencies and percentages, and comparisons between groups were performed using the *Chi-squared test* or *Fisher’s exact test*, as appropriate. To assess agreement between categorical variables, particularly in organism classification by microbiologists versus investigators, *Cohen’s Kappa statistic* was calculated. The strength of associations was further evaluated using effect size metrics such as *Cramer’s V, Bias-corrected Cramer’s V*, and *point-biserial correlation* for appropriate pairings. A p-value of <0.05 was considered statistically significant for all tests. All analyses were conducted using validated statistical software.

## Results

A total of 1600 blood cultures were sent and 205 blood culture-positive episodes were included in the study and evaluated using the structured seven-step clinical-pathological algorithm (Supplementary).

The study included 205 patients with a mean age of 46.19 ± 15.54 years (median: 47, range: 18–80), comprising 92 males (44.9%) and 113 females (55.1%). Most patients were from Uttarakhand (49.8%) and Uttar Pradesh (46.3%), with key contributing districts including Tehri Garhwal (31.7%), Muzaffarnagar (19.8%), and Dehradun (23.8%). Patients were treated in the General Medicine Ward (35.1%), Medicine ICU (40.5%), Pulmonary Medicine (16.1%), and Nephrology Ward (8.3%). (Table No.1)

**Table 1.** Comparison of demographic, clinical, and microbiological characteristics between pathogenic and non-pathogenic isolates.

|  |  |  |  |  |  |
| --- | --- | --- | --- | --- | --- |
| Category | Parameter | Total (n =205) | Non-Pathogen (n=45) | Pathogen (n=160) | p-value |
| Demographic Variables | Age (Years) | 46.19 ± 15.54 | 44.96 ± 14.68 | 46.53 ± 15.80 | 0.534<sup>1</sup> |
|  | Male | 92 (44.8%) | 16 (35.6%) | 76 (47.5%) | 0.155<sup>2</sup> |
|  | Female | 113 (46.2%) | 29 (64.4%) | 84 (52.5%) |  |
|  | Tehri Garhwal district of Uttarakhand | 32 (15.6%) | 0 (0.0%) | 32 (20.0%) | 0.005<sup>3</sup> |
| Site of clinical care |  |  |  |  | 0.007<sup>2</sup> |
|  | General Medicine Ward | 72 (35.1%) | 25 (55.6%) | 47 (29.4%) |  |
|  | Medicine ICU | 83 (40.5%) | 15 (33.3%) | 68 (42.5%) |  |
|  | Nephrology Ward | 17 (8.3%) | 1 (2.2%) | 16 (10.0%) |  |
|  | Pulmonary Medicine | 33 (16.1%) | 4 (8.9%) | 29 (18.1%) |  |
| Microbiological Variables | Klebsiella pneumoniae | 41 (20.0%) | 1 (2.2%) | 40 (25.0%) | <0.001<sup>2</sup> |
|  | Acinetobacter baumannii | 19 (9.3%) | 0 (0.0%) | 19 (11.9%) | 0.016<sup>3</sup> |
|  | Pseudomonas Aeruginosa | 13 (6.3%) | 2 (4.4%) | 11 (6.9%) | 0.738<sup>3</sup> |
|  | CONS | 38 (18.5%) | 19 (42.2%) | 19 (11.9%) | <0.001<sup>2</sup> |
|  | Stenotrophomonas maltophilia | 17 (8.3%) | 10 (22.2%) | 7 (4.4%) | <0.001<sup>3</sup> |
|  | Time to Positivity (hrs) | 17.42 | 21.47 ± 10.13 | 16.29 ± 8.04 | <0.001<sup>4</sup> |
|  | Paired Culture Sent | 103 (50.2%) | 13 (28.9%) | 90 (56.2%) | 0.001<sup>2</sup> |
|  | Both cultures Positive for the Same Organism | 81 (39.5%) | 0 (0.0%) | 81 (50.6%) | <0.001<sup>2</sup> |
|  | Same Organism Blood & Local Site | 57 (27.8%) | 0 (0.0%) | 57 (35.6%) | <0.001<sup>2</sup> |
|  | Catheter Site Isolation | 21 (10.2%) | 0 | 21 (36.8%) | <0.001<sup>2</sup> |
| Clinical Variables | Localized Signs of Infection | 185 (90.2%) | 34 (75.6%) | 151 (94.4%) | <0.001<sup>3</sup> |
|  | SOFA Score within 48 hrs ≥ 2 | 102 (49.7%) | 17 (37.8%) | 85 (53.1%) | 0.069<sup>2</sup> |
|  | Fever | 125 (60.9%) | 16 (47.1%) | 109 (72.2%) | 0.005<sup>2</sup> |
|  | Tachycardia | 68 (33.1%) | 13 (38.2%) | 55 (36.4%) | 0.843<sup>2</sup> |
|  | Tachypnoea | 80 (39.0%) | 15 (44.1%) | 65 (43.0%) | 0.909<sup>2</sup> |
| Localization of Site of Infection | Lower Respiratory Tract | 116 (56.6%) | 21 (46.7%) | 95 (59.4%) | 0.129<sup>2</sup> |
|  | Urinary Tract | 13 (6.3%) | 4 (8.9%) | 9 (5.6%) | 0.488<sup>3</sup> |
|  | Skin and Soft Tissue Infection | 8 (3.9%) | 2 (4.4%) | 6 (3.8%) | 1.000<sup>3</sup> |
|  | CRBSI | 47 (22.9%) | 0 | 47 (29.3%) | <0.001 |
| Complications | Acute liver injury | 36 (17.6%) | 4 (8.9%) | 32 (20.0%) | 0.084<sup>2</sup> |
|  | Acute kidney injury | 26 (12.7 %) | 4 (8.9%) | 22<br>(13.8%) | 0.387<sup>2</sup> |
|  | Haemodialysis | 33 (16.1%) | 5 (11.1%) | 28<br>(17.5%) | 0.303<sup>2</sup> |
|  | Disseminated intravascular<br>coagulation | 15 (16.1%) | 1 (2.2%) | 14 (8.8%) | 0.199<sup>3</sup> |
|  | Septic shock | 47 (22.9%) | 0 (0%) | 47<br>(29.4%) | <0.00<br>1 |
| Outcome | **Death** | 59 (28.8%) | 7 (15.6%) | 52<br>(32.5%) | 0.042 |

**\*\*\*Significant at $p < 0.05$ , 1: t-test, 2: Chi-Squared Test, 3: Fisher's Exact Test, 4: Wilcoxon-Mann-Whitney U Test**

Comorbidities were common among the study population. Hypertension (43.4%) and diabetes mellitus (41.0%) were the most frequent, followed by chronic kidney disease (32.0%) and chronic obstructive pulmonary disease (15.6%). A subset of patients had underlying hepatic or cardiovascular disorders, including chronic liver disease (5.7%) and coronary artery disease (4.9%). Nineteen patients (16.1%) were on maintenance haemodialysis. Other comorbid conditions included hepatitis C infection (9.0%), malignancy (4.1%), hypothyroidism (3.3%), and hepatitis B infection (1.6%).

Single-organism isolates were found in 195 cases (95.1%). The most common organisms were *Klebsiella pneumoniae* (20.0%), CONS (18.5%), *Acinetobacter baumannii* (9.3%), *Stenotrophomonas maltophilia* (8.3%), and *Escherichia coli* (5.4%). The mean time to culture positivity was 17.42 ± 8.78 hours. Paired cultures were submitted in 50.2% of cases, with concordant local-site isolation in 27.8%.

Clinically, 90.2% had signs of localized infection. Common symptoms included fever (67.6%), oxygen requirement (44.3%), tachypnoea (43.2%), and tachycardia (36.8%). SOFA ≥2 occurred in 49.8%. The mean age was comparable between pathogen (46.5 ± 15.8 years) and non-pathogen groups (45.0 ± 14.7 years; p = 0.534). Males were more represented in the pathogen group (47.5%) than in non-pathogens (35.6%), though not statistically significant (p = 0.155). A significantly higher proportion of patients from the Tehri Garhwal district of Uttarakhand belonged to the pathogen group (32/160; 20.0%, p = 0.005). Treatment location also showed a significant association (p = 0.007), with ICU admissions more frequent in the pathogen group (68/160; 42.5%) and general medicine wards in the non-pathogen group (25/45; 55.6%).

*Klebsiella pneumoniae* (25.0%) and *Acinetobacter baumannii* (11.9%) were significantly more common in the pathogen group (p < 0.001 and p = 0.016), while *Coagulase-negative staphylococci* (42.2%) and *Stenotrophomonas maltophilia* (22.2%) predominated among non-pathogens (both p < 0.001). Time to culture positivity was shorter in pathogens (16.3 ± 8.0 hours vs. 21.5 ± 10.1 hours; p < 0.001). Concordant paired cultures and same-site organism isolation occurred exclusively in the pathogen group (p < 0.001).

Clinically, localized infection signs were more frequent in pathogens (94.4% vs. 75.6%; p < 0.001), as was fever (72.2% vs. 47.1%; p = 0.005), other symptoms were not statistically significant. SOFA score ≥2 was more common among pathogens (53.1% vs. 37.8%), though not statistically significant (p = 0.069). (Table No.1)

In the present study, concordant isolation of the same organism from both blood and a corresponding local site was observed in 57 out of 205 patients (27.8%) occurred only in the pathogen group, reinforcing clinical relevance (p < 0.001). Among these 57 cases, the most frequent site of secondary isolation was the respiratory tract, accounting for 27 cases (13.17%), followed by central venous catheter site isolation, 21 (10.2%), and urine culture, 7 (3.4%).

Based on the clinical-microbiological algorithm, the agreement between the clinician and microbiologist in characterizing the organism’s pathogenicity showed that 38 cases (18.5%) were jointly classified as non-pathogenic and 134 cases (65.4%) as pathogenic. The overall raw agreement was 65.4%, with a Cohen’s kappa of 0.25, p < 0.001, indicating fair inter-rater agreement. Discordance was noted in 7 cases (3.4%) where the clinician labeled the isolate non-pathogenic, but the microbiologist considered it pathogenic, and 26 cases (12.7%) with the opposite interpretation. 47 cases (22.9%) classified as pathogenic colonizers. Treating team and the investigator disagreed in 2 (1.5%) cases (1 case was candida isolated from blood which was treated with oral fluconazole for 14 days and another enterococcus faecium which was treated with Injection vancomycin for 2 days which was stopped after 2 days of treatment considering contaminants), which were classified as non-pathogenic as per the investigator and pathogenic as per the treating team.

Clinical outcomes were compared between the pathogen and non-pathogen groups.

Outcomes based on pathogenicity and treatment response demonstrated clear divergence between groups. In the pathogen group (N = 160), 99 patients (61.9%) showed clinical improvement with treatment, while 61 patients (38.1%) did not respond to therapy. Importantly, 60 out of 61 non-responders (98.3.0%) succumbed to their illness, accounting for the majority of deaths in the cohort, and 1 (1.7%) non-responder was admitted at 28 days of follow-up… In contrast, among the non-pathogen group (N = 45), 42 patients (93.3%) were not treated and remained stable, while 3 patients (6.7%) were treated and remained stable. In the non-pathogen group, among patients who did not receive targeted antimicrobial therapy, a 15% mortality was observed. These deaths were attributed to other sources of infection or underlying comorbidities, rather than the cultured organisms themselves. These findings emphasize that non-pathogenic isolates often did not require treatment and were associated with favourable outcomes, whereas a significant proportion of pathogenic isolates were associated with treatment failure and high mortality.

## Discussion

The study demonstrates that combining microbiological data with clinical indicators, such as localized symptoms, SOFA score dynamics, therapeutic response, and source identification, alongside outcome-based follow-up, can markedly enhance the accuracy of pathogen identification. Hence, when both microbiologist and clinician talk in a structured stepwise manner, the best outcomes occur with respect to identifying the exact pathogenicity of an isolated organism in the blood. Several studies have examined blood culture contamination rates; however, most rely solely on microbiological criteria. When clinical features are incorporated, they are often subjective and lack prospective follow-up.(9–12) This study employed a structured stepwise model with a 28-day follow-up to systematically classify blood culture isolates.

Among 1600 blood cultures processed, 205 were culture-positive and included in the analysis. Using the stepwise model, 22% of isolates were classified as non-pathogenic. The overall contamination rate was 2.8%, aligning with prior studies reporting non-pathogen rates ranging from 20% to 56%, depending on clinical context and population. The contamination rate in our study falls within the acceptable range recommended by the American Society for Microbiology (<3%), though a <1% benchmark is ideal.(1,13) The structured algorithm allowed for more nuanced interpretation of isolate significance, improving diagnostic clarity. Its validity was supported by a strong agreement with microbiologist classification (Cohen’s kappa = 0.813, p = <0.001), and treating team classification (Cohen’s Kappa = 0.958, p = <0.001), underscoring its utility in real-world clinical decision-making.

Among the isolated organisms, the most frequent pathogen was *Klebsiella pneumoniae* (n = 41, 20.0%), followed by *Acinetobacter baumannii* (n = 19, 9.3%) and *Pseudomonas aeruginosa* (n = 13, 6.3%). In contrast, coagulase-negative staphylococci (CONS) (n = 38, 18.5%) and *Stenotrophomonas maltophilia* (n = 17, 8.3%) were predominant among non-pathogens. CONS was significantly more common among non-pathogens than pathogens (42.2% vs. 11.9%, p < 0.001). However, 19 CONS isolates (11.9% of pathogens) were classified as pathogenic based on predefined criteria: dual positive blood cultures (n = 5), catheter-related bloodstream infection (CRBSI; n = 9), a ≥2-point rise in SOFA score within 48 hours (n = 2), or new-onset septic shock after 48 hours (n = 3), assessed over a 28-day follow-up. Similarly, *S. maltophilia* was categorized as a pathogen in 7 cases (4.4%) based on consistent clinical symptoms and SOFA score elevation ≥2 in 3 cases. Several studies have identified coagulase-negative staphylococci (CONS) as contaminants, accounting for up to 70% of all contaminated blood cultures, often without incorporating clinical correlation.

However, studies that included clinical context have recognized CONS as a potential pathogen, particularly in cases of catheter-related bloodstream infections (CRBSI). Few studies have attempted clinical correlation; however, most have relied solely on components of the Systemic Inflammatory Response Syndrome (SIRS), often lacking longitudinal follow-up and objective measures such as the Sequential Organ Failure Assessment (SOFA) score.(9,10,14–16).

Maffezzoli et.al, have described the diagnostic and prognostic value of Time to Positivity (TTP), demonstrating that shorter TTP strongly correlates with higher microbial burden, increased severity, and worse outcomes. Patients with Gram-negative bacteraemia (especially *E. coli, Klebsiella pneumoniae*) exhibited median TTP <12 hours, while fungal pathogens such as *Candida spp*. had TTP >36–48 hours, aligning with delayed growth kinetics. The study found that mortality at 28 days was significantly higher in patients with TTP <12 hours, particularly with *K. pneumoniae* and *Pseudomonas aeruginosa*, reinforcing its role as a surrogate marker for critical illness and sepsis severity. Importantly, TTP also helped differentiate true pathogens from contaminants; isolates with TTP >48 hours and lacking clinical correlation were more likely to be coagulase-negative staphylococci or skin flora, thus avoiding unnecessary treatment.(17) Among the 205-blood culture-positive samples in our study, 25 fungal isolates (including *Candida* spp. n = 24 and *Cryptococcus* n =1) were identified. The mean time to positivity (TTP) for bacterial pathogens (n = 142) was 15.01 hours (median: 14, mode: 16, range: 4–44 hours). The mean time to positivity in fungal species was 26.33 hours, and a range of 56 hours (10–66 hours). When all pathogenic organisms were considered, including fungi, the overall mean TTP was 16.29 ± 8.04 hours; excluding fungi, the bacterial pathogen TTP averaged 14.64 hours (range 4-28 hours).

Our study demonstrated that paired culture sampling, concordant isolation from local sites, and clinical symptomatology were valuable indicators of pathogenicity. Paired blood cultures were submitted in 103 cases (50.2%), of which 81 (50.6% of the pathogen group) showed the same organism in both samples, with none in the non-pathogen group (p < 0.001).

Furthermore, the same organism isolated from both blood and a corresponding local site was noted in 57 cases (27.8%), again exclusive to the pathogen group, highlighting the relevance of corroborative culture evidence. Among these, respiratory tract samples accounted for 27 cases (13.17%), catheter tip cultures for 21 (10.2%), and urine cultures for 7 (3.4%), emphasizing that respiratory infections were the most common source of bloodstream seeding in our cohort. In alignment with previous studies, healthcare-associated pneumonia (21%) and CRBSI (21%) remain one of the most common sources of bloodstream infections and is frequently associated with multidrug-resistant pathogens, contributing to prolonged hospitalization and increased mortality.(18)

Clinically, signs of localized infection were more prevalent in the pathogen group (94.4%) than in the non-pathogen group (75.6%; p < 0.001). Fever was also significantly more common in the pathogen group (72.2% vs. 47.1%, p = 0.005). However, other clinical variables such as tachycardia, tachypnoea, and elevated SOFA scores showed trends but did not reach statistical significance. Nonetheless, these findings affirm that systemic inflammatory response and organ dysfunction, when integrated with microbiological findings, can aid in distinguishing clinically significant infections.

This study highlights a clear distinction in clinical outcomes based on the pathogenicity of blood culture isolates. Among patients with pathogenic organisms, over one-third (38.1%) failed to respond to treatment, with an alarmingly high mortality rate of 98.3% in this subgroup, underscoring the strong association between treatment failure and poor prognosis. In contrast, those who responded to treatment demonstrated significantly better outcomes. Notably, three pathogen-positive patients who were treated remained clinically stable, suggesting a possible window for early therapeutic success even in uncertain cases.

In the non-pathogen group, none received targeted antimicrobial therapy, yet 93.3% remained clinically stable without intervention. A small proportion (6.7%) were treated and remained stable, indicating cautious overtreatment. Although a 15% mortality rate was observed in the non-pathogen group, these deaths were attributed to other causes and not the cultured organisms. These findings reinforce the clinical value of accurate pathogen classification and support the judicious use of antimicrobials in culture-positive patients.

Population-based studies have shown that bloodstream infections (BSIs) are associated with a 30-day mortality of approximately 25%, with the most implicated pathogens being *Escherichia coli, Klebsiella* spp., *Pseudomonas aeruginosa*, and *Staphylococcus aureus*.(19)

One key limitation of this study is the inherent subjectivity involved in classifying organisms as pathogenic or non-pathogenic, despite the use of a structured stepwise model. Inter-observer variability between clinician and microbiologist interpretations may have influenced the categorization, particularly for organisms commonly regarded as commensals or contaminants (e.g., CONS or *Stenotrophomonas*). Moreover, since the primary investigator was also a clinician involved in patient care, there exists a potential bias toward the treating physician’s clinical judgment, which may have affected the objectivity of classification before microbiological review. Although the stepwise model incorporates predefined checklists for consistency, the sequential flow of these steps has not yet been validated through empirical evidence and remains based largely on expert consensus. Finally, this was a single-center study with a relatively short follow-up period (up to 28 days), which may limit the generalizability of the findings and the ability to assess long-term outcomes or late complications.

## Conclusion

In this prospective study of 205 blood culture-positive patients, application of a structured seven-step clinical-pathological model enabled more accurate classification of pathogens and non-pathogens. Of the total isolates, 160 (78.0%) were determined to be pathogenic and 45 (22.0%) non-pathogenic. Klebsiella pneumoniae (20.0%), Acinetobacter baumannii (9.3%), and Pseudomonas aeruginosa (6.3%) were the most frequently identified pathogens, while CONS (18.5%) and Stenotrophomonas maltophilia (8.3%) were predominant among non-pathogens. The mean TTP was significantly shorter among pathogens (16.3 ± 8.0 h) compared to non-pathogens (21.5 ± 10.1 h), with bacterial pathogen TTP averaging 15.01 hours. Paired culture concordance and same-site isolation occurred exclusively in the pathogen group (p < 0.001). Respiratory tract infections (59.4 %) and CRBSI (29 %) were the leading sources of bloodstream infections in pathogen group.

Agreement with microbiologists (Cohen’s kappa = 0.813) and treating clinicians (κ = 0.958) further validated the robustness of the algorithm. This model, combining clinical, microbiological, and follow-up data, offers a pragmatic framework for improving diagnostic stewardship, guiding treatment decisions, and optimizing patient outcomes in resource-limited and high-burden settings.

## Data Availability

All data produced in the present study are available upon reasonable request to the authors

## Acknowledgment

None

## Authors’ contributions

All authors made substantial contributions to conception and design, acquisition of data, or analysis and interpretation of data; took part in drafting the article or revising it critically for important intellectual content; agreed to submit to the current journal; gave final approval of the version to be published; and agree to be accountable for all aspects of the work.

